# Viability-Compatible Preservation for Mass Cytometry Using Cisplatin Pulse Quenching and Gradual Formaldehyde Release

**DOI:** 10.1101/2025.08.08.25333023

**Authors:** Leonie Wagner, Christopher Mark Skopnik, Diana Metzke, Paul Freund, Franca Alena Hegemann, Joram Arzig, Pouneh Mirkheshti, Nina Goerlich, Jan Klocke, Philipp Enghard, Sabine Baumgart

## Abstract

**Objective:** Mass cytometry (CyTOF) enables high-dimensional single-cell profiling, but especially for non-blood derived samples relies on immediate processing, limiting its application to fragile, low-yield clinical specimens such as urine. We developed a novel preservation protocol that combines viability staining with gentle fixation, enabling cryopreservation and delayed processing.

**Methods:** Building on a previously established protocol using imidazolidinyl urea (IU) and MOPS buffer for flow cytometric applications, we enhanced the protocol for CyTOF by incorporating a one-minute pulse of cisplatin (5□µM) for live–dead discrimination. A novel quenching step with DL-methionine (5□mM) was introduced to minimize background signal without compromising antigen integrity. Samples were fixed overnight at 4□°C and cryopreserved prior to CyTOF analysis. The protocol was validated using peripheral blood mononuclear cells (PBMCs) and was applied to urine samples.

**Results:** The preservation method maintained single-cell integrity, surface marker expression, and cisplatin staining specificity. DL-methionine efficiently quenched residual cisplatin reactivity. The protocol performed robustly across a wide range of cell inputs and maintained consistency following freeze–thaw cycles. In urine samples, immune cell subset frequencies and viability discrimination were comparable between the optimized and standard staining protocols.

**Conclusion:** Our protocol enables delayed processing and cryopreservation of fragile clinical samples while preserving compatibility with mass cytometry workflows. By integrating cisplatin-based viability staining, DL-methionine quenching, and slow-release fixation, this method supports standardized immune profiling of low-abundance samples in clinical and translational research.

## INTRODUCTION

Mass cytometry (cytometry by time-of-flight, CyTOF) has emerged as a powerful platform for high-dimensional immune profiling, enabling the simultaneous detection of over 40 distinct proteins at the single-cell level. The technology combines principles of flow cytometry and mass spectrometry-based detection of metal isotopes conjugated to antibodies.^1^ The quality and interpretability of CyTOF data strongly depend on effective sample preservation, particularly when immediate processing is not feasible. Proper preservation allows batch processing of large sample cohorts and stabilizes the cellular state at a defined time point for downstream analysis. This includes maintaining the activation status, cell morphology, and antigenic structures.

Several studies have demonstrated the utility of SmartTube Proteomic Stabilizer (PROT1) for preserving both surface and intracellular epitopes in whole blood samples.^2-4^ However, the stabilization of non-blood-derived cell types such as tissue-resident or tumor-infiltrating cells, and particularly urinary cells remains challenging. These cells are often exposed to harsh, non-physiological conditions characterized by nutrient deprivation, osmotic stress, and toxic metabolites, which can rapidly compromise cellular integrity.

To address this limitation, our group introduced a two-step conservation protocol for flow cytometry using imidazolidinyl urea (IU), a formaldehyde-releasing agent, in combination with MOPS buffer in prior works.^5^ This method effectively preserved urinary cell morphology and antigen integrity for delayed flow cytometric analysis. It was successfully applied in the assessment of various inflammatory kidney diseases, kidney transplant rejection or acute kidney injury.^6-8^

Recently, we used mass cytometry for deep phenotyping of urinary leukocytes in freshly collected urine samples from active lupus nephritis patients, demonstrating its potential as a non-invasive tool for immune monitoring.^9^ While the analytical capabilities of CyTOF are undisputed, its dependence on freshly processed samples remains a significant logistical challenge.

In this study, we propose a novel preservation protocol for mass cytometry combining a slow-release fixation process with a pulse viability staining. This approach is based on our previously published IU-MOPS fixation method^5^ and standardized cisplatin viability staining^10^ modified by a new quenching agent. Our data show the maintenance of cellular structures, minimization of background signal, and preservation of epitope integrity, thereby enabling reliable mass cytometric analysis of fragile clinical samples such as urine. It expands the applicability of CyTOF in both a clinical and translational research setting.

## METHODS

### Ethics

The study was conducted in accordance with the ethical principles outlined in the Declaration of Helsinki and by our institution. Ethical approval was granted by the Ethics Committee of Charité University Hospital (EA1/284/19). Written informed consent was obtained from all participants prior to their enrollment in the study.

### Sample collection

For all preliminary experiments, freshly collected human heparin-anticoagulated whole blood was subjected to red blood cell lysis (Erylysis buffer, Qiagen) in order to obtain human peripheral blood leukocytes (or white blood cells, WBC). Peripheral blood mononuclear cells (PBMCs) were generated according to standard protocol using density gradient centrifugation with Ficoll (Cytiva, Marlborough, Massachusetts, USA).^11^ For generating dead cells, PBMCs were incubated with 4% para-formaldehyde (PFA, BD Cytofix Fixation Buffer, BD Biosciences, New Jersey, USA) for 30 min at room temperature (RT). We collected 10 urine samples of kidney graft recipients, kidney graft recipients, who shed a heterogeneous spectrum of immune cells in relatively high numbers into the urine. Samples were obtained as first morning urine or, whenever possible, via urinary catheter, and transported directly to the lab on ice for further processing.

### Cell preservation protocol including viability staining for CyTOF measurement

#### Optimized protocol

For our preservation technique we applied our previously published protocol using imidazolidinyl urea (IU, Sigma-Aldrich Chemie GmbH, Taufkirchen, Germany) and MOPS (Carl Roth GmbH & Co. KG, Karlsruhe, Germany), hereafter referred to as IUM, to dilute and buffer predominantly non-physiological biological samples such as urine.^5^ To adapt the procedure for CyTOF analysis, we incorporated cisplatin (CP) (cis-platinum (II)-diamine dichloride; Enzo Life Sciences GmbH, Lörrach, Germany) at a final concentration of 5□µM for live–dead cell discrimination as determined by prior titration experiments (data not shown), confirming the protocol described by *Fienberg et al*.^10^ The one minute pulse reaction was quenched by adding DL-methionine (Sigma-Aldrich Chemie GmbH, Taufkirchen, Germany) at an optimal final concentration of 5 mM (see results section). After 24 h incubation at 4°C, samples were centrifuged, washed with phosphate buffered saline (PBS, Deutsches Rheuma-Forschungszentrum Berlin (DRFZ)) supplemented with 0.5 % bovine serum albumin (BSA, PAN Biotech, Aidenbach, Germany) and 0.02% sodium azide (Sigma-Aldrich Chemie GmbH, Taufkirchen, Germany) (PBS/BSA) once and cryopreserved in fetal calf serum/dimethylsulfoxid (FCS/DMSO, 90%/10%, DRFZ) at -80°C (Fig. 1A).

**Fig. 1.**
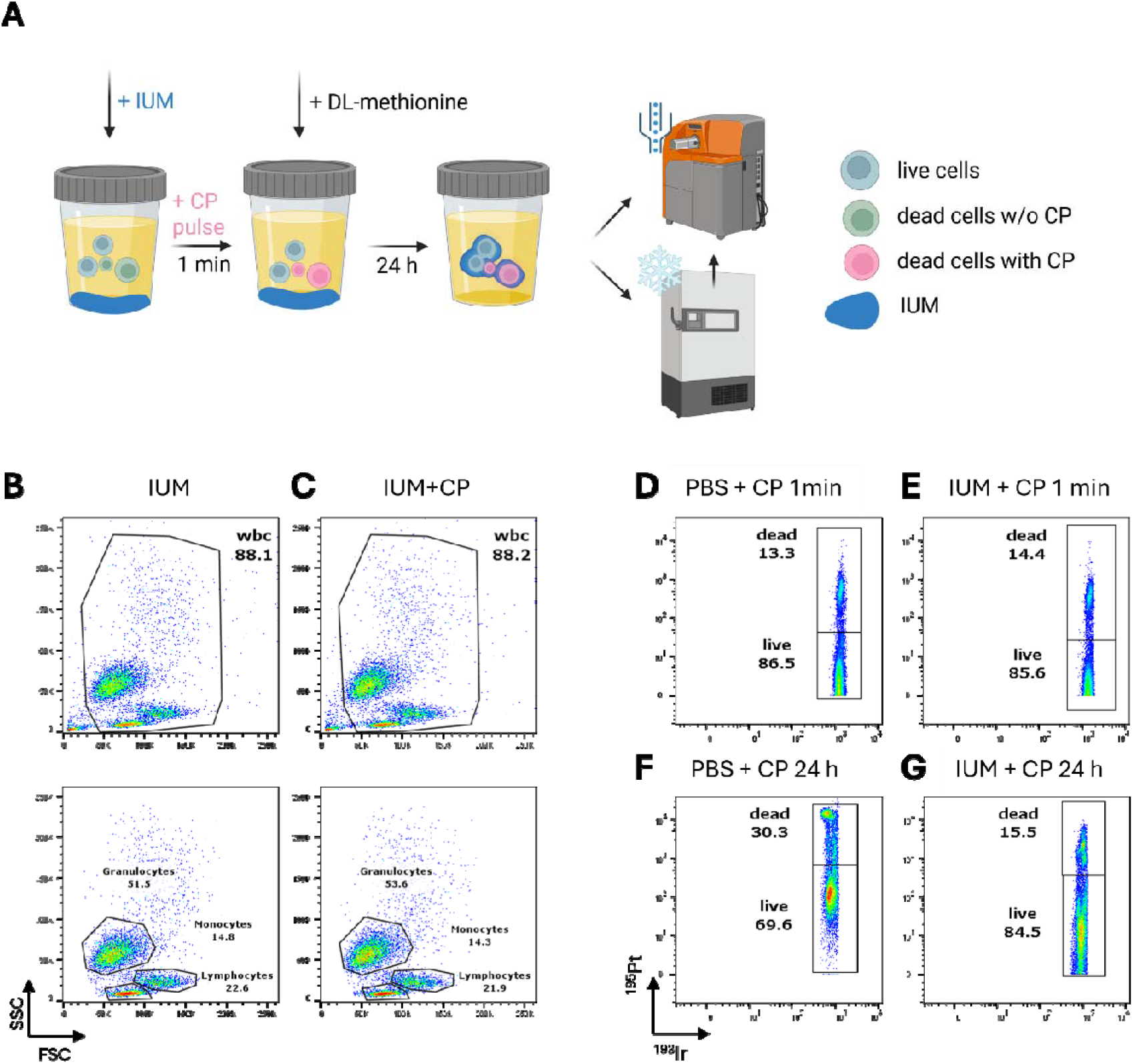
Integration of cisplatin (CP) viability staining into a slow-release fixation protocol. **(A)** Workflow for viability staining integrated in a slow-release fixation process for mass cytometry (MC) analysis. **(B-C)** Interaction of viability staining and fixation method using peripheral blood samples - FC scatter parameter plots with frequencies of main leukocyte populations after incubation overnight with **(B)** IUM only and **(C)** IUM and CP. **(D-G)** Interaction of viability staining and fixation method using PBMCs - MC plots with frequencies of live and dead cells: **(D)** CP in PBS pulsed for one minute. **(E)** CP in IUM pulsed for one minute. **(F)** CP in PBS for 24 h. **(G)** CP in IUM for 24 h. Samples were immediately processed and measured after indicated incubation time. Gates were adjusted for MMI. CP, cisplatin; IUM, imidazolidinyl urea in MOPS buffer; MMI, mean metal intensity; wbc, white blood cells. Fig 1A created with BioRender.com.

Deviating parameters during method development are described in detail in the results section.

#### Standard protocol for CP staining

For comparison, we used a standard protocol for CP staining: Cells were stained with CP at a final concentration of 5 µM in PBS for one min and immediately quenched by PBS/BSA. Samples were washed twice with PBS/BSA and further processed for the subsequent CyTOF measurement according to the OMIP-34 protocol.^12^

### Sample preparation for CyTOF and measurement

Frozen samples were thawed, filtered through a 30 µm mesh and washed with PBS/BSA twice, fresh samples were directly processed according to the OMIP-34 protocol.^12^ Briefly, cells were treated with Beriglobin as FcR-blocking reagent before staining with our antibody panel (Table 1). Cells were then washed, fixated with 1.6% PFA and incubated with Iridium-intercalator (Standard BioTools (SBT), South San Francisco, USA) in permeabilization buffer (Life Technologies GmbH, Darmstadt, Germany) for 1 h at RT. Following this, two wash cycles with PBS/BSA were conducted, followed by one cycle with Millipore Q water. The cell suspension was supplemented with 10% v/v Four Element Beads (SBT). Final cell concentration was adjusted to 7.5 × 10^5^ cells/mL before measurement. A Helios instrument (SBT) operated with the instrument control software v6.7.1014 was set up according to the manufacturer instructions. Data was normalized before further processing.

**Table 1:**
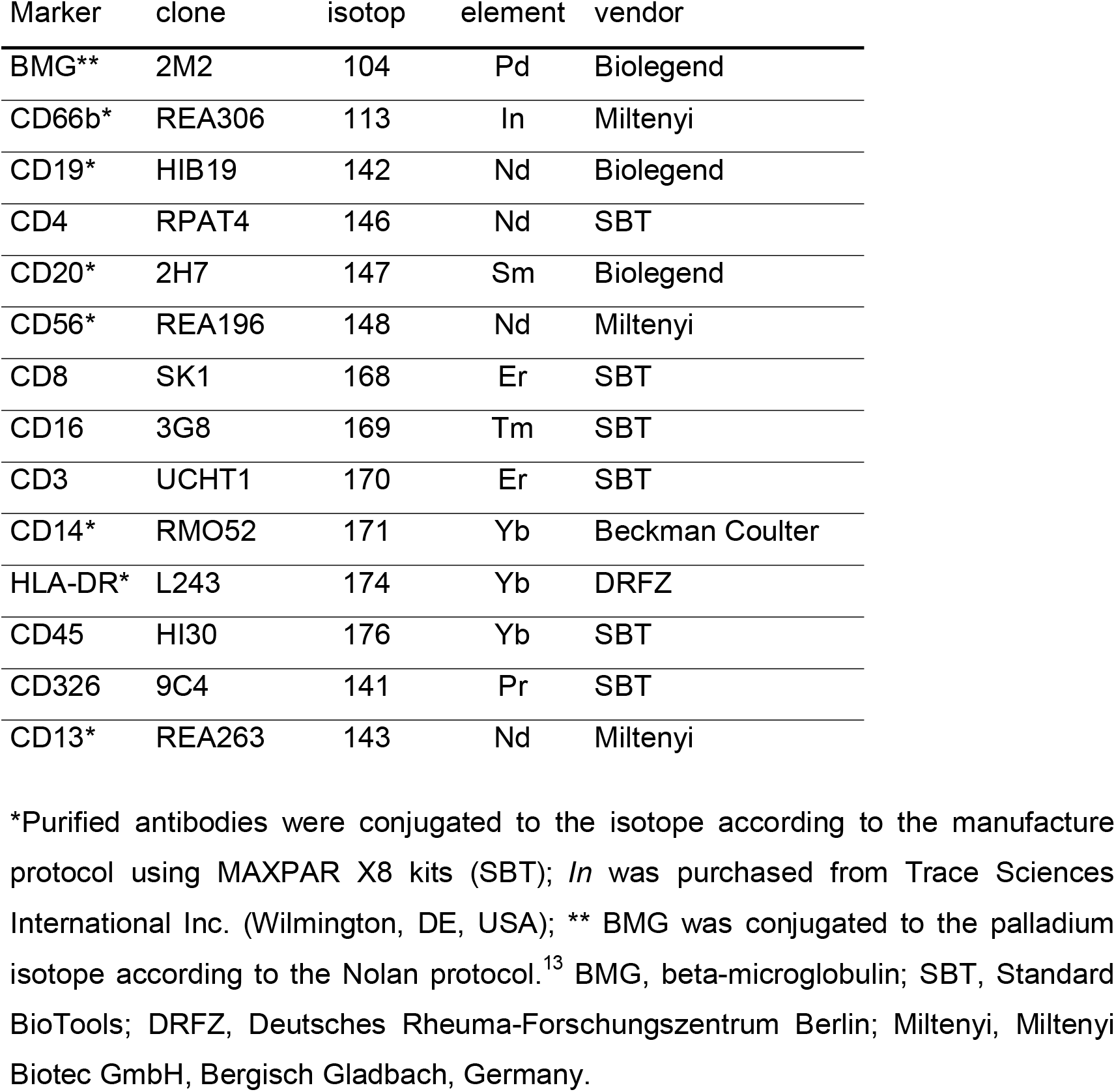
CyTOF antibody panel

| Marker | clone | isotop | element | vendor |
| --- | --- | --- | --- | --- |
| BMG** | 2M2 | 104 | Pd | Biolegend |
| CD66b* | REA306 | 113 | In | Miltenyi |
| CD19* | HIB19 | 142 | Nd | Biolegend |
| CD4 | RPAT4 | 146 | Nd | SBT |
| CD20* | 2H7 | 147 | Sm | Biolegend |
| CD56* | REA196 | 148 | Nd | Miltenyi |
| CD8 | SK1 | 168 | Er | SBT |
| CD16 | 3G8 | 169 | Tm | SBT |
| CD3 | UCHT1 | 170 | Er | SBT |
| CD14* | RMO52 | 171 | Yb | Beckman Coulter |
| HLA-DR* | L243 | 174 | Yb | DRFZ |
| CD45 | HI30 | 176 | Yb | SBT |
| CD326 | 9C4 | 141 | Pr | SBT |
| CD13* | REA263 | 143 | Nd | Miltenyi |

\*Purified antibodies were conjugated to the isotope according to the manufacture protocol using MAXPAR X8 kits (SBT); *In* was purchased from Trace Sciences International Inc. (Wilmington, DE, USA); \*\* BMG was conjugated to the palladium isotope according to the Nolan protocol.<sup>13</sup> BMG, beta-microglobulin; SBT, Standard BioTools; DRFZ, Deutsches Rheuma-Forschungszentrum Berlin; Miltenyi, Miltenyi Biotec GmbH, Bergisch Gladbach, Germany.

### Quality check of urine samples using flow cytometry (FC)

For the FC quality check, urine was centrifuged at 600 x G for 8 min and washed in PBS/BSA with 2mM Ethylen diamain tetraacecetic acid (EDTA, Invitrogen, California, USA) (PBE). One aliquot was used as unstained control, the other was incubated with FcR-blocking reagent (Miltenyi Biotec GmbH, Bergisch Gladbach, Germany) and stained with the following antibodies: anti-CD3 (APC efluor780, SK7, eBioscience, San Diego, CA, USA), anti-CD4 (PE-Vio770, REA623, Miltenyi), anti-CD8 (Alexa647, GN11/134D7, DRFZ), anti-CD14 (FITC, Tük4, Miltenyi), anti-CD45-BUV805 (BD, HI30, New Jersey, USA), DAPI (DRFZ). After incubation for 20 min at 4 °C, samples were washed and resuspended in PBE to be measured at a FACS Canto II (BD Biosciences, San Jose, CA, USA). FC controls were introduced to serve as a screening tool for urine sample quality before sample processing for mass cytometry.

### Statistical analysis and data plotting

Data was analyzed using the FlowJo software (v. 10.8., FlowJo, BD Biosciences). Statistical analysis was carried out using R Studio^14^ (e4392fc9, 2024-06-05), specifically the following packages: tidyverse (v. 2.0.0), ggpubr (v 0.6.0), fcexpr (v. 0.0.0.9975).

## RESULTS

### Preservation of structural integrity of cells after addition of cisplatin to the slow-release fixation process

In order to evaluate the feasibility of adding cisplatin (CP) along with the fixative IUM, we first investigated cell size- and granularity-related flow cytometric parameters (side scatter (SSC), forward scatter (FSC)).

Human peripheral blood leukocytes were incubated overnight with IUM only or IUM and CP, washed and resuspended in Millipore Q water the following day. FC measurements show similar frequencies of the main leukocyte populations in both the IUM and IUM+CP fixated samples, respectively (Fig. 1B,C). Comparable overall cell counts in both conditions indicate that no significant cell loss occurred. Furthermore, the absence of cell aggregates or debris in scatter plots supports the preservation of single-cell integrity throughout the fixation process.

### Compatibility of live-dead cell discrimination using cisplatin in the slow-release fixation process

Next, we tested CPs capability of labeling dead cells in the context of IUM fixation in comparison to the standard protocol for CP staining.

Here, a mixture containing viable and dead PBMCs was incubated with IUM and 5 µM CP for 1 min and 24 h. As a control, a similar set of cells was incubated with CP only in a non-fixating buffer (PBS) using same durations and temperature (Fig. 1D-G).

There was no difference in the frequency of dead cells (^195^Pt^+^ cells) when comparing conditions with and without fixating buffer within 1 min CP pulse reaction (Fig. 1D,E). After 24 h incubation of CP in IUM, frequencies of dead cells were still comparable (Fig. 1G). However, the mean metal intensity (MMI) for both the CP-negative (^195^Pt^-^) and –positive (^195^Pt^+^) population was increased along with the extended incubation (Fig. 1F,G). This phenomenon was more pronounced in the absence of fixation (Fig. 1F), suggesting that IUM not only has fixative properties but also exerts a quenching effect on the CP reaction.

### Introduction of DL-methionine as a quenching enhancer for CP reaction

Minimizing exposure time to CP is crucial to prevent accidental labeling of viable cells due to its slow diffusion. While *Fienberg et al*.^10^ used RPMI/FCS to stop the CP reaction, we tested DL-methionine, a sulfur-containing amino acid, for its quenching properties, following a similar approach (Fig. 2). Freshly prepared PBMCs were stained with the optimized 5□µM cisplatin concentration in IUM for one minute, after which DL-methionine was added at concentrations ranging from 5□µM to 50□mM for titration. (Fig.□2A). The optimal concentration of 5□mM DL-methionine was identified, which efficiently quenched the CP reaction. We could show that the MMI of CP was significantly reduced when using DL-methionine compared to its absence (Fig. 2 B-D).

**Fig. 2.**
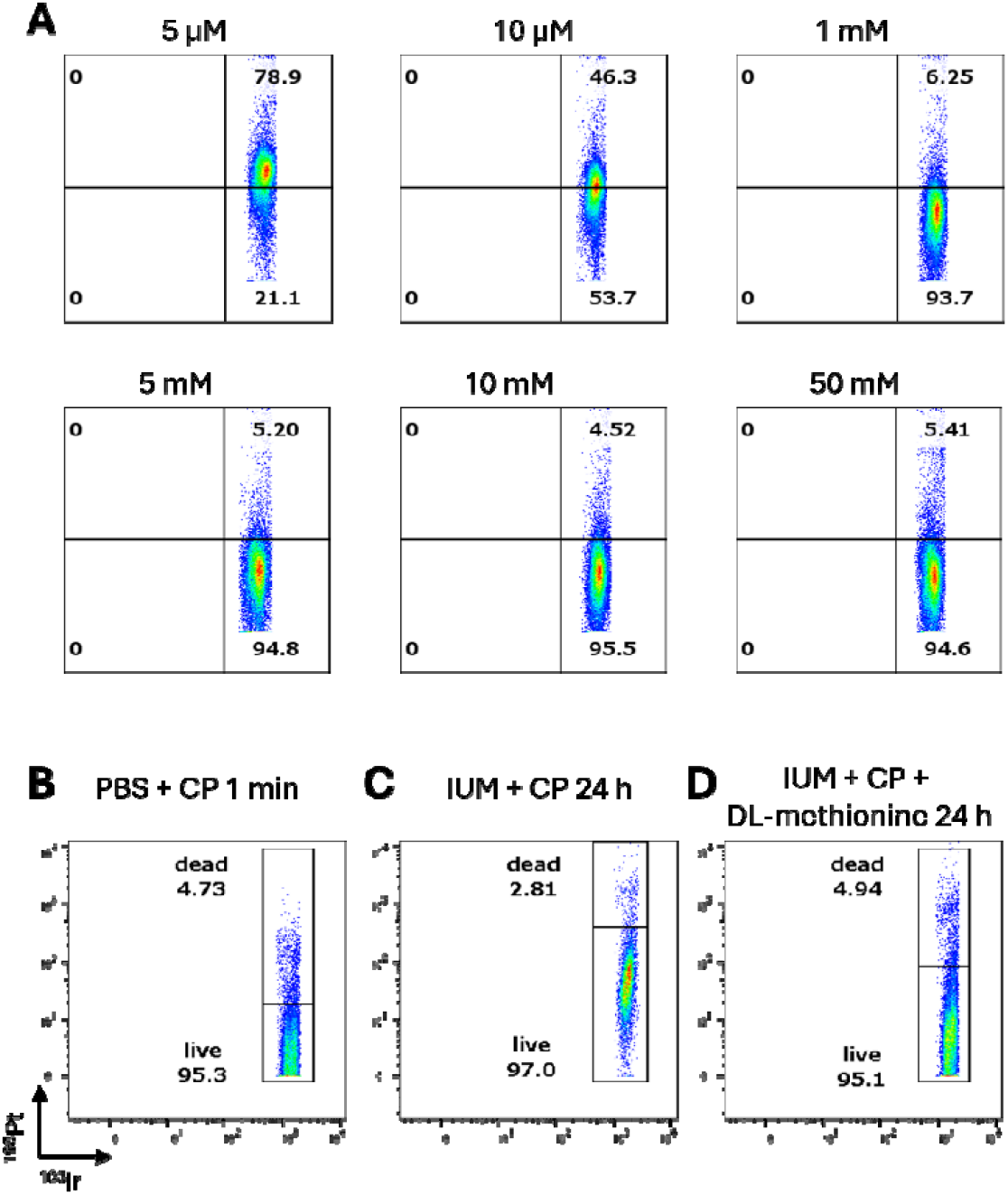
Introduction of DL-methionine as an effective quenching agent to stop the CP reaction. **(A)** Freshly prepared PBMCs in IUM were stained for 1 min with CP. A serial dilution of DL-Methionine (5□µM to 50□mM) was tested to identify the optimal final concentration required to efficiently quench the CP pulse reaction. **(B-D)** Reduction of MMI using DL-methionine comparing **(B)** standard protocol with **(C)** IUM + CP for 24 h without an additional quenching agent and **(D)** optimized protocol with DL-methionine as quenching enhancer. CP, cisplatin; IUM, imidazolidinyl urea in MOPS buffer, MMI, mean metal intensity.

### Maintenance of CP specificity under slow-release fixation conditions

An important factor that needed ruling out was the prolonged staining of fixated and thus dead cells. To investigate the specificity of CP staining, we compared two conditions: The standard protocol involving a 1 min CP pulse on dead PBMCs spiked with a portion of live CD45 (^176^Yb)-labeled PBMCs under non-fixed conditions (Fig 3A), and the same sample analyzed in 24 h IUM (Fig. 3B). There were no significant differences in the frequencies of dead cells (^195^Pt^+^/^176^Yb^-^) and live cells (^195^Pt^-^/^176^Yb^+^). The consistent low frequency of approximately 3% of double-positive cells (^195^Pt^+^/^176^Yb^+^) indicated that with our protocol, CP retained high specificity, even after a 24 h incubation, when a substantial portion of the fixation reagent is expected to have been released and exerted its effect.

**Fig. 3.**
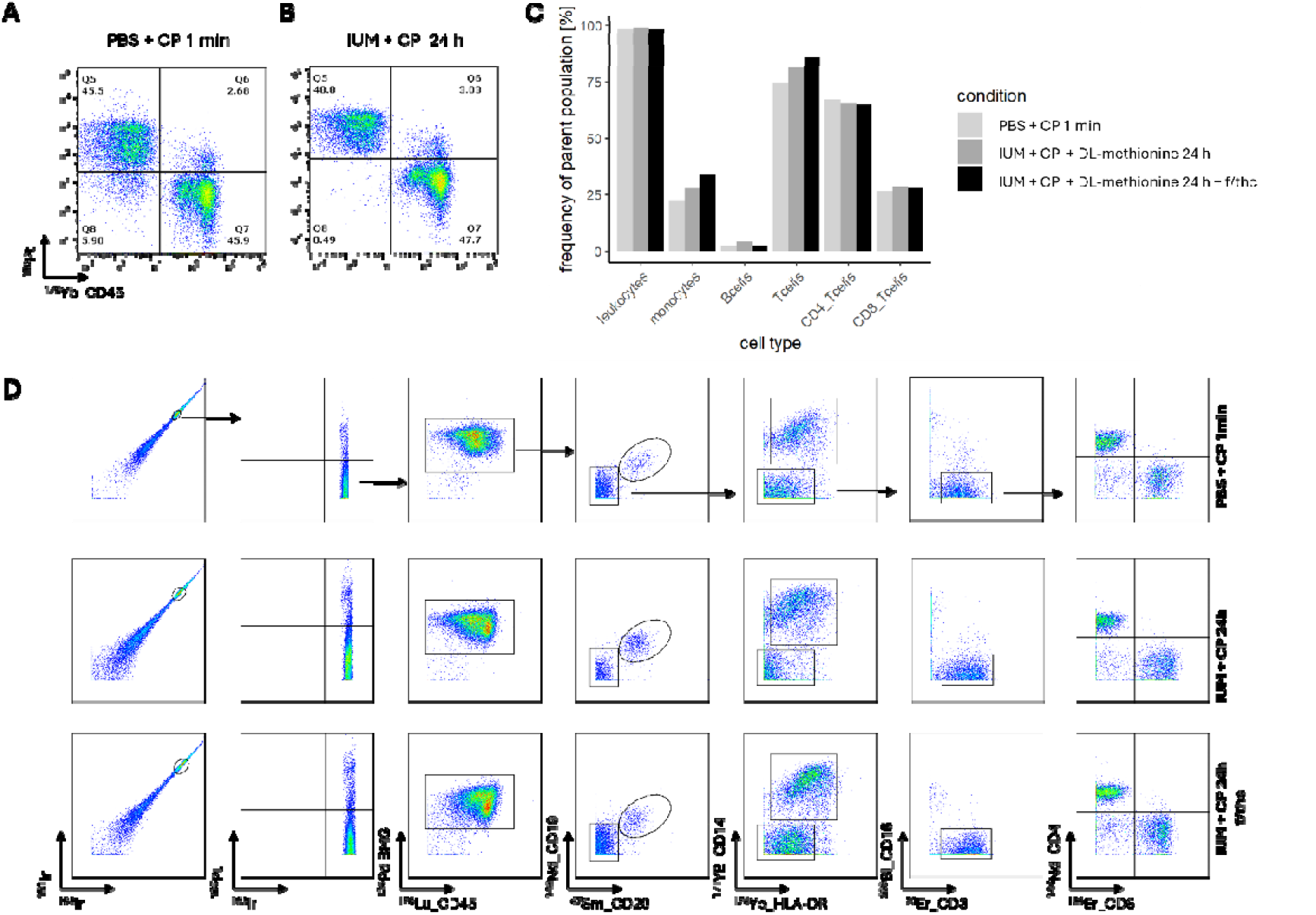
Specific overnight staining of dead PBMCs and preserved antigen integrity. **(A-B)** A mixture of viable PBMCs that were labeled with CD45 (^176^Yb) and PFA-fixated PBMCs (dead cells) were stained with CP in **(A)** PBS for 1 min using standard protocol **(B)** in IUM under fixating conditions incubated overnight. Gates were adjusted to account for increased MMI. **(C)** Quantification of population frequencies following different sample treatments: PBMCs treated with CP in PBS for 1 minute (standard), IUM with CP and DL-methionine for 24□h, and IUM with CP and DL-methionine for 24□h including one f/thc. (**D**) Exemplary gating strategy for selected populations across the three tested conditions, with each row representing one condition: After excluding internal standard beads (not shown), doublets, and dead cells (^195^Pt^+^), total leukocytes were gated based on BMG and CD45. B cells were defined as CD19^+^CD20^+^. Monocytes were identified as CD14^+^HLA-DR^+^ after B cell exclusion. Remaining CD3^+^CD16^−^ cells were classified as T cells and further subdivided into CD4^+^ and CD8^+^ subsets. Staining remains specific and comparable across treatments. PBMCs, peripheral blood mononuclear cells; PFA, para-formaldehyde; CP, Cisplatin; IUM, imidazolidinyl urea in MOPS buffer; MMI, mean metal intensity; f/thc, freeze–thaw cycle.

### Preservation of antigen integrity after fixation process including viability staining/quenching and cryopreservation

The performance of the optimized protocol compared to the standard protocol was first tested on PBMCs. Additionally, we examined the impact of cryopreservation following the 24 hour fixation to assess its applicability for multi-center studies or batch processing. We applied our optimized protocol, in which a one minute CP pulse for live-dead discrimination was quenched using DL-methionine, embedded within a slow-release fixation process, to PBMCs first. This was combined with antibody staining to analyze the relative frequencies of major immune cell subsets; For exemplary dot plots and gating strategy detailing our antibody panel, see Fig. 3D. Notably, we observed that a freeze/thaw cycle (f/tc) reduced CP-associated background signal. Importantly, the consistent frequencies of key immune cell subsets across all tested conditions and workflows highlight both the robustness of the protocol and its compatibility with cryopreservation (Fig. 3C, D).

### Staining efficiency of CP independent on cell numbers

Our aim was to apply the optimized protocol to patient urine samples. Based on prior knowledge, we anticipated a wide variation in cell numbers across patient samples, potentially spanning several orders of magnitude.^9^ To account for this, we performed a titration of cell input using a defined mixture of 50% viable and 50% dead PBMCs across a range from 1□×□10□ to 1□×□10□ cells (Supp. Fig. 1).

In a subset of samples, a freeze–thaw cycle was introduced to mimic potential sample handling conditions. No significant differences in the frequency of dead cells were observed between samples subjected to a freeze–thaw cycle and those that were not. (Supp. Fig. 1).

### Application of the optimized method to patient urine samples

Subsequently, we applied our optimized method to urine samples followed by MC analysis. Urine samples were split and processed under two different conditions: one aliquot was analyzed immediately following a 1 min CP incubation (standard protocol), while the other was subjected to our optimized protocol involving CP pulse reaction, DL-methionine quenching in IUM, and a 24 h slow-release fixation (Fig. 4A-C). Dead cell frequencies did not differ significantly when comparing both conditions (Fig. 4A,B). However, median metal intensity (MMI) for cisplatin was higher in urine samples compared to PBMCs, likely due to the presence of urinary epithelial cells.

**Fig. 4.**
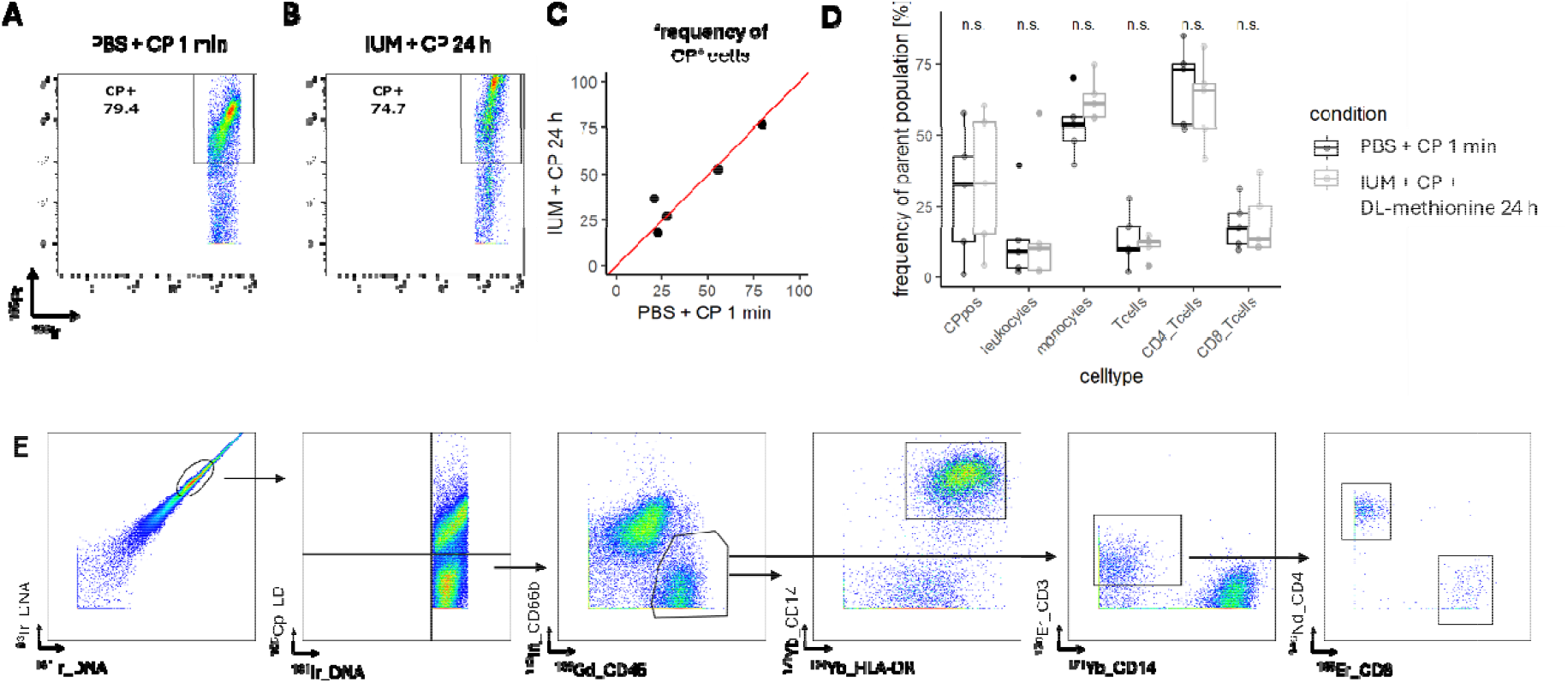
Application of the optimized protocol to patient urine samples. **(A-B)** Comparison of CP MMI in **(A)** 1 min CP staining in PBS under non-fixated conditions (standard protocol) and **(B)** 1 min CP/quenched by methionine in IUM under fixated conditions for 24 h (optimized protocol). **(C)** Frequencies of dead cells (CP^+^) of both conditions as described in A and B from 5 kidney transplant urine samples. **(D)** No significant difference in frequencies of major leukocyte subsets in urine after treatment with standard vs. optimized protocol. **(E)** Exemplary dot plots and gating strategy: After excluding internal standard beads (not shown), doublets, and dead cells (^195^Pt^+^), granulocytes were excluded (CD45^dim^CD66b^+^). Remaining CD45^+^ cells were further classified as monocytes (CD14^+^HLADR^+^) and T cells (CD3^+^CD14^-^).T cells were further subdivided into CD4^+^ and CD8^+^ subsets. CP, cisplatin; MMI, mean metal intensity; IUM, imidazolidinyl urea in MOPS buffer.

Finally, we measured urine samples from ten patients (Fig. 4C-E). To ensure sufficient cell counts for mass cytometry analysis, samples containing fewer than 1,000 CD45^+^ cells per 100 mL were excluded, resulting in five samples in the final evaluation. Exemplarily MC plots are illustrated in the Fig. 4E. Fresh urine samples processed using the standard protocol showed no significant differences in urinary cell subset frequencies compared to those treated with our optimized slow-release fixation method, confirming previous results that fixation, including CP staining and quenching, does not affect staining quality or antigen integrity (Fig. 4D).

## DISCUSSION

In this study, we present a novel preservation protocol for mass cytometry, building upon the previously established flow cytometry method by *Freund et al*., which employs imidazolidinyl urea (IU) in MOPS buffer to enable gentle, slow-release fixation. We adapted this protocol for compatibility with the CyTOF platform by integrating a cisplatin-based viability staining approach commonly used in mass cytometry. Notably, we modified the original method introduced by *Fienberg et al*. through the implementation of a novel quenching reagent, thereby ensuring compatibility with the fixation process and preserving sample integrity for downstream analyses. This methodological advancement significantly extends the utility of CyTOF in clinical and translational research settings, particularly for analyzing fragile or low-abundance samples such as urine, which often contain a high and variable proportion of dead cells.^9^

This approach addresses a critical limitation in mass cytometry workflows—the requirement for immediate sample processing—which can pose significant logistical challenges, especially in clinical and multi-center studies. By enabling cryopreservation of overnight-fixated cells, our protocol permits delayed processing and long-term storage. This flexibility supports batch analysis and promotes standardized immune profiling across diverse research and clinical settings.

A key finding of our work is the consistency of detection of important immune cell subsets under different sample conditions. In both PBMCs and urine samples, our protocol resulted in comparable frequencies of CD45^+^-associated cell populations as in freshly measured controls. This indicates reliable preservation of cellular surface markers that is a crucial criterion for the interpretation of high-dimensional cytometry data.

The rapid uptake of cisplatin (CP) by cells with compromised membranes, combined with its high reactivity toward protein nucleophiles and the formation of stable covalent bonds, makes CP an ideal reagent for live-dead cell discrimination under conditions of slow-release fixation. Cisplatin’s protein-targeting mechanism enables robust labeling, outperforming traditional mass cytometry viability dyes such as rhodium (Rh)- or iridium-based intercalators^15^ or Rh(III)-loaded DOTA-NHS esters.^1^

Due to its covalent binding properties, cisplatin offers superior stability across multiple washing steps, which is particularly advantageous in workflows involving cryopreservation. Our observation that urine samples exhibit a generally higher cisplatin signal compared to PBMCs is likely attributable to the greater abundance of non-immunological cell types, such as renal epithelial and urothelial cells, which are often already damaged or exhibit apoptotic features.

To prevent unintended labeling of viable cells due to slow diffusion, it is critical to minimize the exposure time to CP. In the standard CP staining protocol, RPMI supplemented with fetal calf serum (FCS) is used as a quenching agent. The high protein content in FCS can compete with cellular proteins for binding to fixatives, particularly under slow-release fixation conditions, thereby potentially reducing fixation efficiency. For this reason, neither FCS nor PBS/BSA was suitable for stopping the CP reaction in our protocol. Interestingly, when using IUM alone (without additives), we observed already a minimal quenching effect of the reaction.

To ensure complete termination of the CP reaction, we sought a cost-effective, non-toxic, and user-friendly alternative that would not interfere with the fixation process. Methionine, a naturally occurring amino acid, has been shown to readily form covalent adducts with cisplatin, effectively blocking its interaction with canonical targets such as DNA. This quenching effect has been demonstrated both in vitro and in vivo.^16,17^

Leveraging these properties, we employed methionine as a quenching agent in our protocol. Our results demonstrate that methionine effectively quenches cisplatin activity even under slow-release fixation conditions, without interfering with immune cell staining or viability discrimination.

While the protocol performed well across multiple immune cell markers, potential limitations include its effect on sensitive epitopes such as chemokines and phospho-proteins, which require further investigation. Additionally, the relatively low cell yields particular in some urine samples may limit applicability for the mass cytometry approach due to the extended washing steps compared to flow cytometry, underscoring the need for careful sample quality control.

Overall, our preservation strategy significantly expands the usability of mass cytometry for liquid biopsies, offering a practical solution for high-dimensional immune monitoring in translational and clinical research.

## Conclusion

In summary, we established a robust and practical preservation protocol for mass cytometry that enables the high-dimensional analysis of clinical samples such as urine, even after delayed processing and cryopreservation. By incorporating a brief cisplatin-based viability stain followed by effective quenching using DL methionin at the start of an overnight slow-release fixation process, our method ensures epitope preservation, minimizes background, and maintains cellular integrity. This approach facilitates standardized workflows, supports batch processing, and expands the feasibility of CyTOF in multi-center and translational research settings.

## Data Availability

All data produced in the present study are available upon reasonable request to the authors.

## Author Contributions

SB, PE and CMS designed the study. DM, CMS and SB conducted experiments. PF and NG recruited participants. CMS, NG, DM and LW analyzed data. LW, SB, JK and PE wrote, LW, SB, NG, JK, PE, PM, JA, CMS and FAH edited the manuscript.

## Acknowledgments

We thank all study participants for their participation. Part of this research was supported by the BIH Ad-hoc Booster grant awarded to NG. We also acknowledge the Flow Cytometry and Cell Sorting Facility and the laboratory managers at the Deutsches Rheuma-Forschungszentrum Berlin for their technical expertise and insightful support. In addition, we are grateful to the staff of the nephrology and intensive care wards at Charité for their assistance with sample collection.

## References

1. Bendall SC, Simonds EF, Qiu P, Amir el AD, Krutzik PO, Finck R, Bruggner RV, Melamed R, Trejo A, Ornatsky OI, Balderas RS, Plevritis SK, Sachs K, Pe’er D, Tanner SD, Nolan GP. Single-cell mass cytometry of differential immune and drug responses across a human hematopoietic continuum. Science. 2011;332(6030):687–696.

2. Geanon D, Lee B, Gonzalez-Kozlova E, Kelly G, Handler D, Upadhyaya B, Leech J, De Real RM, Herbinet M, Magen A, Del Valle D, Charney A, Kim-Schulze S, Gnjatic S, Merad M, Rahman AH. A streamlined whole blood CyTOF workflow defines a circulating immune cell signature of COVID-19. Cytometry Part A. 2021;99(5):446–461.

3. Rahil Z, Leylek R, Schürch CM, Chen H, Bjornson-Hooper Z, Christensen SR, Gherardini PF, Bhate SS, Spitzer MH, Fragiadakis GK, Mukherjee N, Kim N, Jiang S, Yo J, Gaudilliere B, Affrime M, Bock B, Hensley SE, Idoyaga J, Aghaeepour N, Kim K, Nolan GP, McIlwain DR. Landscape of coordinated immune responses to H1N1 challenge in humans. J Clin Invest. 2020;130(11):5800–5816.

4. Smets T, Stevenaert F, Adams HC, 3rd, Vanhoof G. Deep Profiling of the Immune System of Multiple Myeloma Patients Using Cytometry by Time-of-Flight (CyTOF). Methods in molecular biology (Clifton, NJ). 2018;1792:47–54.

5. Freund P, Skopnik CM, Metzke D, Goerlich N, Klocke J, Grothgar E, Prskalo L, Hiepe F, Enghard P. Addition of formaldehyde releaser imidazolidinyl urea and MOPS buffer to urine samples enables delayed processing for flow cytometric analysis of urinary cells: A simple, two step conservation method of urinary cells for flow cytometry. Cytometry Part B, Clinical cytometry. 2023;104(6):417–425.

6. Goerlich N, Brand HA, Langhans V, Tesch S, Schachtner T, Koch B, Paliege A, Schneider W, Grützkau A, Reinke P, Enghard P. Kidney transplant monitoring by urinary flow cytometry: Biomarker combination of T cells, renal tubular epithelial cells, and podocalyxin-positive cells detects rejection. Sci Rep. 2020;10(1):796.

7. Prskalo L, Skopnik CM, Goerlich N, Freund P, Wagner L, Grothgar E, Mirkheshti P, Klocke J, Sonnemann J, Metzke D, Schneider U, Hiepe F, Eckardt KU, Salama AD, Bieringer M, Schreiber A, Enghard P. Urinary CD4+ T cells Predict Renal Relapse in ANCA-Associated Vasculitis: Results of the PRE-FLARED Study. Journal of the American Society of Nephrology : JASN. 2024.

8. Grothgar E, Goerlich N, Samans B, Skopnik CM, Metzke D, Klocke J, Prskalo L, Freund P, Wagner L, Duerr M, Matz M, Olek S, Budde K, Paliege A, Enghard P. Urinary CD8+HLA-DR+ T Cell Abundance Non-invasively Predicts Kidney Transplant Rejection. Frontiers in medicine. 2022;9:928516.

9. Bertolo M, Baumgart S, Durek P, Peddinghaus A, Mei H, Rose T, Enghard P, Grutzkau A. Deep Phenotyping of Urinary Leukocytes by Mass Cytometry Reveals a Leukocyte Signature for Early and Non-Invasive Prediction of Response to Treatment in Active Lupus Nephritis. Front Immunol. 2020;11:256.

10. Fienberg HG, Simonds EF, Fantl WJ, Nolan GP, Bodenmiller B. A platinum-based covalent viability reagent for single-cell mass cytometry. Cytometry Part A : the journal of the International Society for Analytical Cytology. 2012;81(6):467–475.

11. Noble PB, Cutts JH, Carroll KK. Ficoll Flotation for the Separation of Blood Leukocyte Types. Blood. 1968;31(1):66–73.

12. Baumgart S, Peddinghaus A, Schulte-Wrede U, Mei HE, Grützkau A. OMIP-034: Comprehensive immune phenotyping of human peripheral leukocytes by mass cytometry for monitoring immunomodulatory therapies. Cytometry Part A. 2017;91(1):34–38.

13. Han G, Spitzer MH, Bendall SC, Fantl WJ, Nolan GP. Metal-isotope-tagged monoclonal antibodies for high-dimensional mass cytometry. Nature Protocols. 2018;13(10):2121–2148.

14. Team R. RStudio: Integrated Development for R. RStudio. PBC, Boston, MA. 2022.

15. Ornatsky OI, Lou X, Nitz M, Schäfer S, Sheldrick WS, Baranov VI, Bandura DR, Tanner SD. Study of cell antigens and intracellular DNA by identification of element-containing labels and metallointercalators using inductively coupled plasma mass spectrometry. Analytical chemistry. 2008;80(7):2539–2547.

16. Vrana O, Brabec V. L-methionine inhibits reaction of DNA with anticancer cis-diamminedichloroplatinum(II). Biochemistry. 2002;41(36):10994–10999.

17. Sooriyaarachchi M, White WM, Narendran A, Gailer J. Chemoprotection by D-methionine against cisplatin-induced side-effects: insight from in vitro studies using human plasma. Metallomics : integrated biometal science. 2014;6(3):532–541.

