## Supplemental Figure 1 for "Viability-Compatible Preservation for Mass Cytometry Using Cisplatin Pulse Quenching and Gradual Formaldehyde Release"

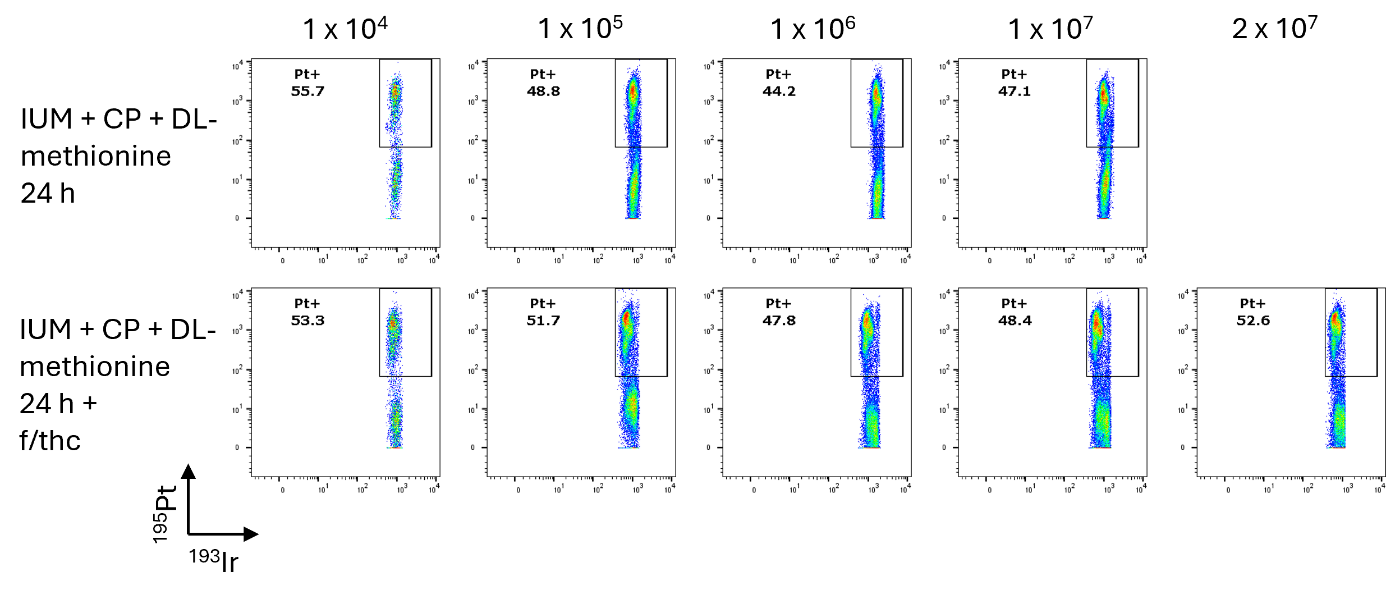

Supplemental Fig.1 **Assessment of protocol robustness across varying cell input and cryopreservation conditions.** Cell input was titrated using a defined mixture of live and dead PBMCs (50%/50%) and processed following our optimized protocol with and without an additional freeze and thaw cycle. Frequencies of dead cells (Pt^+^ cells) are indicated in the plots. PBMCs, peripheral blood mononuclear cells; CP, Cisplatin; IUM, imidazolidinyl urea in MOPS buffer; f/thc, freeze–thaw cycle.
